# Rapid, inexpensive methods for exploring SARS CoV-2 D614G mutation

**DOI:** 10.1101/2021.04.12.21255337

**Authors:** Sirwan M.A. Al-Jaf, Sherko Subhan Niranji, Zana Hameed Mahmood

**Author notes:** Corresponding author: Department of Biology, College of Education, University of Garmian, Berdasur campus, Kalar, 46021, Kurdistan Regional Government, Iraq.

## Abstract

A common mutation has occurred in the spike protein of severe acute respiratory syndrome coronavirus 2 (SARS CoV-2), known as D614G (A23403G). There are discrepancies in impacting of this mutation on the virus’s infectivity, and the whole genome sequencings are expensive and time-consuming. This study aims to develop three fast economical assays for prompt identifications of the D614G mutation including Taqman probe-based real-time reverse transcriptase polymerase chain reaction (rRT PCR), an amplification refractory mutation system (ARMS) RT and restriction fragment length polymorphism (RFLP), in nasopharyngeal swab samples. Both rRT and ARMS data showed G614 mutant indicated by presence of HEX probe and 176bp, respectively. Additionally, the results of the RFLP data and DNA sequencings confirmed the prevalence of G614 mutant. These methods will be important, in epidemiological, reinfections and zoonotic aspects, through detecting the G614 mutant in retro-perspective samples to track its origins and future re-emergence of D614 wild type.

## 1. Introduction

Since its first emergence in Wuhan, China in late December 2019, severe acute respiratory syndrome coronavirus 2 (SARS CoV-2) which leads to coronavirus disease 2019 (Covid-19), has been considered as pandemic since March 2020 by World Health Organisation (WHO) and it has caused approximately 107.8 million global infections and 2.37 million deaths by 12 February 2021 (COVID-19 Dashboard by the Centre for Systems Science and Engineering (CSSE) at Johns Hopkins University (JHU) https://coronavirus.jhu.edu/map.html). The genome of SARS CoV-2 contains several genes encoding structural proteins such as spike (S), envelope (E), membrane (M), and nucleocapsid (N) (Ahmadpour et al., 2020). SARS CoV-2 interacts with angiotensin-converting enzyme 2 (ACE2) of human cells through its spike proteins, spike 1 (S1) and spike 2 (S2) (Walls et al., 2020), which are cleaved, by human type II transmembrane serine protease (TMPRSS2) and furin, to facilitate the viral envelope fusion with the targeted human cell membrane (Ahmadpour et al., 2020; Walls et al., 2020). It is worth mentioning that S1 protein has three main domains: C-terminal domain (CTD), receptor-binding domain (RBD) which binds to human ACE2, and N-terminal domain (NTD) (Ahmadpour et al., 2020).

However, SARS CoV-2 has proofread mechanisms for correcting its RNA replication errors, nevertheless, mutations occur in its genome leading to an increase in viral survival adaptations (Pachetti et al., 2020; Phan, 2020; Robson et al., 2020; Romano et al., 2020). One of the most common mutations occurred, since its emergence, has been recognized as D614G (A23403) at amino acid number 614 in the spike protein sequence (nucleotide sequence number 23403) of the reference Wuhan SARS CoV-2 genome, when nucleotide A in G**<u>A</u>**T encoding aspartate residue (D) altered to G creating G**<u>G</u>**T, a codon for glycine (Badua et al., 2020). The D614G mutation is located in the NTD of S1 that lies between RBD and S2 near the cleavage site (Bhattacharyya et al., 2020), where both S1 and S2 are cleaved by TMPRSS2 and furin (Ahmadpour et al., 2020; Walls et al., 2020). Therefore, this mutation may enhance the viral infectivity of SARS CoV-2 by increasing the attachment capability of the RBD to human ACE2 via decreasing interactions between S1 and S2 (Gupta et al., 2020).

Since the emergence of the SARS CoV-2 D614G mutation, tracing back to January 2020 from China to Europe (Xu et al., 2020), and its current highly prevalence in all continents of the world (Bhattacharyya et al., 2020; Gómez-Carballa et al., 2020), there have been controversial studies on impacts of this single nucleotide variation (SNV), D614G, on the viral survival fitness (Isabel et al., 2020; Kim et al., 2020; Omotuyi et al., 2020; Yong Zhang et al., 2020), immunogenicity and antigenic epitopes (Gupta et al., 2020; Hernández-Huerta et al., 2020; Islam et al., 2020; Kim et al., 2020; Koyama et al., 2020; Saha et al., 2020; To et al., 2020), antibody neutralising sensitivity (Garcia-Beltran et al., 2020; Goldman et al., 2020; Hu et al., 2020;Klumpp-Thomas et al., 2020; Li et al., 2020; Mansbach et al., 2020; Plante et al., 2020), infectivity (Daniloski et al., 2020;Hu et al., 2020; Korber et al., 2020; Li et al., 2020), transmission (van Dorp et al., 2020) and fatality (Hernández-Huerta 2020). The studies have mainly focused on bioinformatic simulation models, and sequence alignments comparing with other coronavirus genomes with the available SARS CoV-2 whole viral genome sequences. They predicted spike protein destabilizing effect of the D614G mutation, leading to rapid S1 detachment with S2 causing more ACE2 attachments. Few studies have conducted investigations of D614G mutation on *in vitro* viral pseudotype infected cells (Daniloski et al., 2020; Hou et al., 2020; Hu et al., 2020; Korber et al., 2020; Li et al., 2020; Wang et al., 2020), laboratory animal models (Plante et al., 2020), and infected populations, suggesting increases in infectivity, viral loads (Korber et al., 2020; Yurkovetskiy et al., 2020; L. Zhang et al., 2020), and fatality (Hernández-Huerta et al., 2020). Overall, data indicated the viral adaptability to human cells as a result of D614G mutation. However, few studies have considered this single mutation outside RBD as a consequence of random mutations (Dearlove et al., 2020; van Dorp et al., 2020; Grubaugh et al., 2020; Isabel et al., 2020). Thus, these discrepancies should be elucidated to see whether the mutation is due to viral fitness or a random process (Korber et al., 2020).

On one hand, no adequate cohort clinical data have been obtainable for finding causality or even associations between this mutation and Covid-19 patients’ severity. On the other hand, sufficient whole genome sequences might be available only in countries with highly developed genome services. However, whole genome sequencings are expensive and time-consuming. Moreover, there should have been plethora of SARS CoV-2 positive nasopharyngeal samples stored in developing countries without being sequenced due to lack of whole genome sequence facilities. Thus, retro-perspective studies are required to discover the origin of the virus imported into those countries.

Therefore, developing inexpensive and rapid methods for identifying SNVs, such as D614G, are essential for tracking this variant by epidemiologists, molecular virologists or immunologist collaborating with clinicians to compare Covid-19 patients with SARS CoV-2 D614 and G614 variants. Previous study mistakenly developed an RFLP method for identifying another mutation at residue 615 of the spike protein (Hashemi et al., 2020), but not D614G as commented by Niranji and Al-Jaf (Niranji and Al-Jaf, 2021). Recent study highlighted the necessity of SARS CoV-2 D614G mutant using biosensing and restriction enzyme methods including BtsCI endonuclease which can cleave the wild type D614 but do not cut G614 mutant (Yang Zhang et al., 2020).

Therefore, the purpose of this study is to develop three various methods such as Taqman probe based rRT PCR, ARMS and RFLP to detect SARS CoV-2 D614G mutation in clinical nasal swab samples taken from Covid-19 patients. Furthermore, DNA sequencings were used for approving the validity of the methods.

## 2. Materials and methods

### 2.1. Sample collection and study area

Three (3) ml viral transport medium (VTM) containing nasopharyngeal swab samples were collected in Covid-19 clinically suspected individuals (n=67) at Coronavirus Research and Identification Lab in the University of Garmian in Kalar town, Sulaymaniyah province, Kurdistan region of Iraq from June to October 2020. The VTM samples were preserved on ice or 4°C or -85°C until viral nucleic acids were extracted. Written consent forms were taken from the covid-19 suspected persons and the study was ethically approved by an ethical committee, which is adhered to WHO Guidelines on Ethical Issues in Public Health Surveillance and to the principles of the Declaration of Helsinki, at the department of biology, University of Garmian.

### 2.2. Viral RNA Extraction

Sample processing was handled according to WHO standards under biological safety Level 2 using personal protection equipment (PPE) and biological safety cabinet (Labconco, Kansas City, MO, USA). Total viral RNA was extracted from the VTM preserved nasal swab samples using AddPrep Viral Nucleic Acid Extraction Kit (AddBio, Korea). According to the manufacturer’s instructions, 200 µl of the VTM stored samples mixed with 350 µl lysis buffer and 3.5 µl β-mercaptoethanol (14.2M) in a 1.5 ml microcentrifuge tube. After 10 minutes of incubating the lysed mixture, 150 µl of Isopropanol was added and this was followed by two successive washing steps, using 500 µl washing 1 and 2 solutions in a spin column, centrifuged for 13,000 rpm for 1 minute. Finally, the bound RNA was eluted with 50 µl elution buffer and the purified RNA samples were kept at 4°C for short-term storage or -85°C deep freeze for long period preservations.

### 2.3. Detection of SARS CoV-2 RNA by rRT PCR

The RNA extracted samples were inspected for SARS CoV-2 using genesig® Real-Time PCR assay (PrimerdesignTM Ltd, UK). The assay has been originally designed to be one step rRT PCR for making both cDNA synthesis from viral RNA and amplification of the cDNA using primer-probes, specific for SARS CoV-2 RdRp, developed in a single-step reaction. According to the manufacturer’s instructions, a mixture of 10 µl Oasig™ OneStep 2X RT-qPCR master mix and 2 µl primer-probe mix was prepared. The prepared RT qPCR mixture was mixed with either 8 µl RNA, negative or positive controls, amplified using CFX Connect Real-Time PCR Detection System (Bio-Rad, Germany) at the following amplification programs: reverse transcription at 55°C for 10 minutes, initial denaturation at 95°C for 2 minutes, followed by 50 cycles of denaturation at 95°C for 10 seconds and annealing at 60°C for 60 seconds.

### 2.4. Detection of SARS CoV-2 D614G mutations

SARS CoV-2 RNA samples, identified by Gensig rRT PCR kit, with Cq values <30, were selected for detection of the D614G mutation by using Taqman probe-based rRT PCR, ARMS, and RFLP.

#### 2.4.1. Taqman probe-based rRT PCR

Forward and reverse primers (D614G IN F and D614G IN R), designed to cover both sides of the D614G mutation (A23403G), with a product size of 169 bp, from the Wuhan strain (GenBank: MN908947.3), were carefully inspected using NCBI online database for checking melting temperatures, GC contents, lengths, product sizes and locations of the primers. Taqman probes including D614 D-FAM and G614 G-HEX (Macrogen, South Korea) were designed to be specific for each wildtype Adenine (A) and mutant Guanine (G), respectively, as shown in Table 1 and Figure 1B. The lyophilized primers and probes were reconstituted with RNase/DNase Free Water to 100 µM which was further diluted to 10 µM. Ten (10) µl of Addprobe rRT PCR master mix (Addbio, South Korea) was mixed with 0.5 µl (10 µM) of each primer (D614G IN F and D614G IN R) and probes (D614 D-FAM and G614 G-HEX) followed by adding 8 µl of RNA, giving 20 µl total volume with 250 nM final concentrations of primer probes. The rRT PCR reactions were performed as follow: reverse transcription at 50°C for 20 minutes, initial denaturation at 95°C for 10 minutes, followed by 50 cycles of denaturation at 95°C for 10 seconds, and annealing at 61°C for 60 seconds using CFX Connect Real-Time PCR Detection System (Bio-Rad, Germany).

**Table 1:**
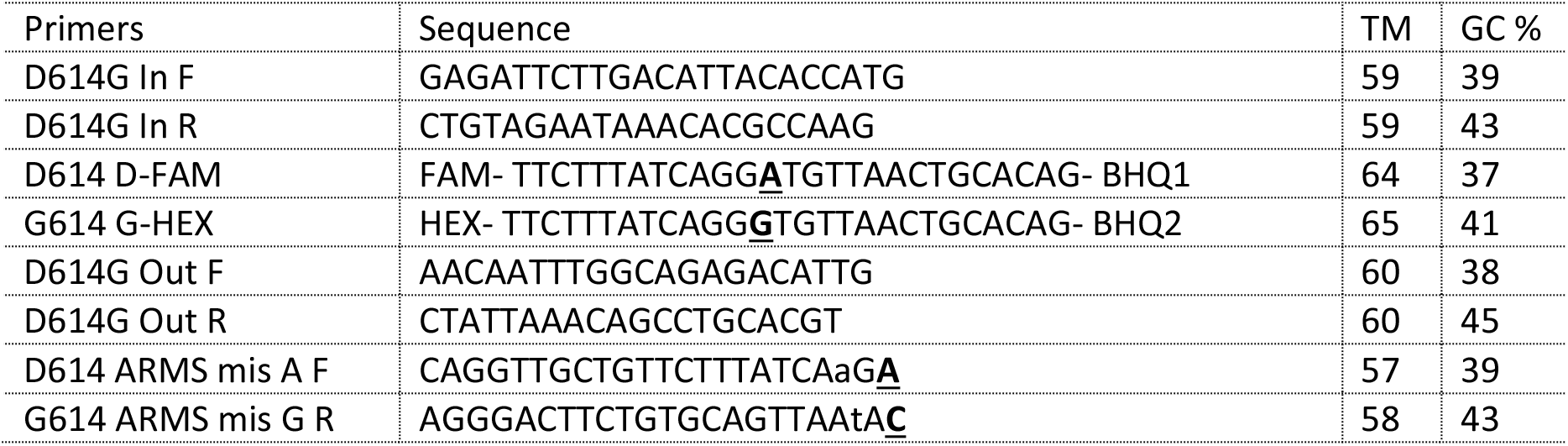
Primers and probes for rRT PCR, ARMS and RFLP

**Figure 1:**
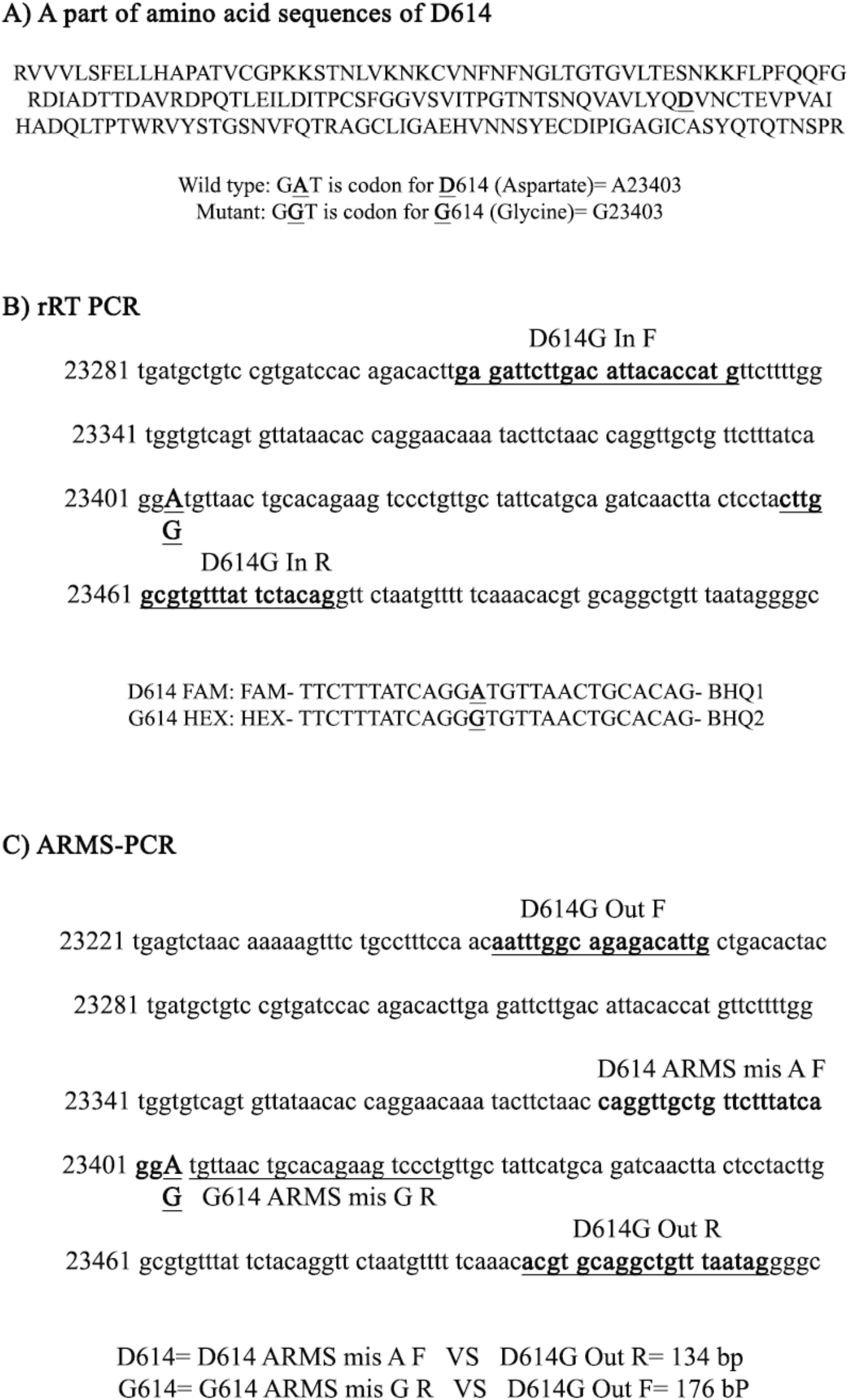
Locations of primers and probes in a specific region around D614G (**A**23403**G**) mutation in SARS CoV-2 isolate Wuhan-Hu-1, complete genome. GenBank: MN908947.3. A part of amino acid sequences showing amino acid D614 which is mutated to G614 (A). Locations of rRT PCR Primers and probes (B). Locations of ARMS PCR primers (C).

#### 2.4.2. Amplification-refractory mutation system PCR (ARMS-PCR)

Four primers were designed and used in a single tube multiplex reaction including D614G outer and ARMS specific primers as shown in Table 1 and Figure 1C. Outer primers, D614G Out F and D614G Out R that amplify a PCR product size of 266 bp in which both A and G variants were located. D614G ARMS specific primers were designed as follows: <u>D</u>614 ARMS **<u>A</u>** F amplifies the wildtype **<u>A</u>** with the D614 Out R creating a 134 bp PCR product size. <u>G</u>614 ARMS **<u>G</u>** R amplifies the mutant **<u>G</u>** with the D614 Out F primer creating a 176 bp PCR product (Table 1 and Figure 1C). ARMS primers were designed in a manner that the wild type **<u>A</u>** nucleotide is located at the 3’end of the forward primer while the mutant **<u>G</u>** nucleotide is located at the 3’end of the reverse primer. To reduce non-specific amplification, a nucleotide mismatch was introduced, just two nucleotides before the 3’end of each ARMS primer. The ARMS PCR reaction mixtures were as follows: 0.5µl of ARMS primers (10µM) added to 10 µl of Addscript RT PCR master mix (Addbio) and 8 µl RNA. The PCR program was set using conventional Lightcycler (Eppendorf, Germany) as follow: reverse transcription at 50°C for 20 minutes, initial denaturation at 98°C for 10 minutes, followed by 36 cycles of denaturation at 95°C for 15 seconds, annealing at 61°C for 45 seconds, and extension 72°C for 30 seconds and then a final extension at 72°C for 5 minutes.

#### 2.4.3. RFLP method for D614G variant

This methods was developed using NEB cutter (https://nc2.neb.com/NEBcutter2/index.php). Interestingly, BtsCI restrictions site (5’-GG**<u>A</u>**TGNN-3’) was found in the wild type D614 (G**<u>A</u>**T) but not in G614 mutant (G**<u>G</u>**T). Thus, this enzyme was used to distinguish between D614 from G614 as the former nucleic acid sequence is cleaved but the latter remains uncut (Figure 2A). Therefore, PCR products, amplified by D614G Out primers, were incubated with BtsCI restriction endonuclease at 50°C for 30 minutes in BtsCI buffer and inactivated at 80°C for 20 minutes as recommended by the manufacturer (New England Biolabs, Ipswich, MA, USA).

**Figure 2:**
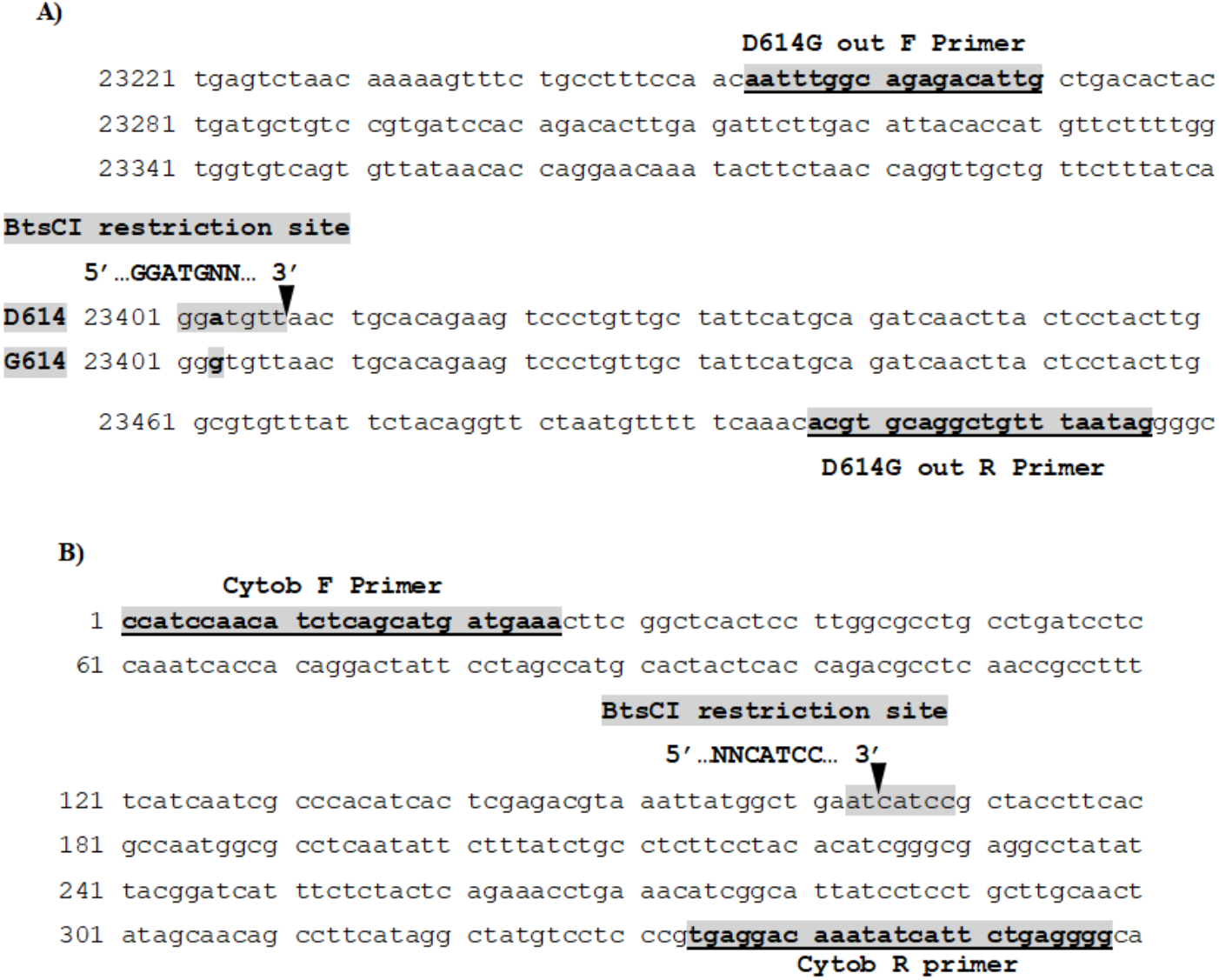
DNA sequencings of both SARS CoV-2 D614G and human cytochrome b used for RFLP: Panel (A) Nucleic acid sequences of SARS CoV-2 shows restriction site (GGATGNN) in the wild type D614 which is cleaved by BtsCI enzyme while G614 mutant sequence is not cut by the enzyme. Both D614G Out F and R primers are also located. Panel B) Nucleic acid sequences of human cytochrome b gene shows restriction site (5’…NNCATCC…3’) which is cleaved by BtsCI enzyme. Both universal F and R primers are also located.

Furthermore, to confirm the action of the enzyme, a positive control was exploited by amplifying a human cytochrome b gene using universal primers (Kocher et al., 1989) since its DNA sequence contains BtsCI restriction site (5’…NNCATCC…3’) using the NEB cutter as shown in Figure 2B.

#### 2.4.4. DNA sequencings

Three (3) PCR products, amplified by conventional RT PCR using D614G Out primers, were randomly sent for Sanger sequencings (Macrogen Co., Seoul, KR), to confirm D614G mutants and the sequences were submitted to NCBI using Bankit (Benson et al., 2015).

## 3. Results

The current study has developed three methods including probe-based rRT PCR, ARMS and RFLP. For the first time, using fast, cost-effective methods, this study has identified G614 mutants in 67 nasopharyngeal samples in Iraq.

### 3.1. Probe-based rRT PCR

In the primer-probe rRT PCR method, two primers (D614G IN forward and reverse) with two probes (D-FAM and G-HEX) were designed for detecting D614 and G614 variants of SARS CoV-2, respectively. The probes and primers worked in 250 nM final concentrations at 61°C as shown in Figure 3.

**Figure 3:**
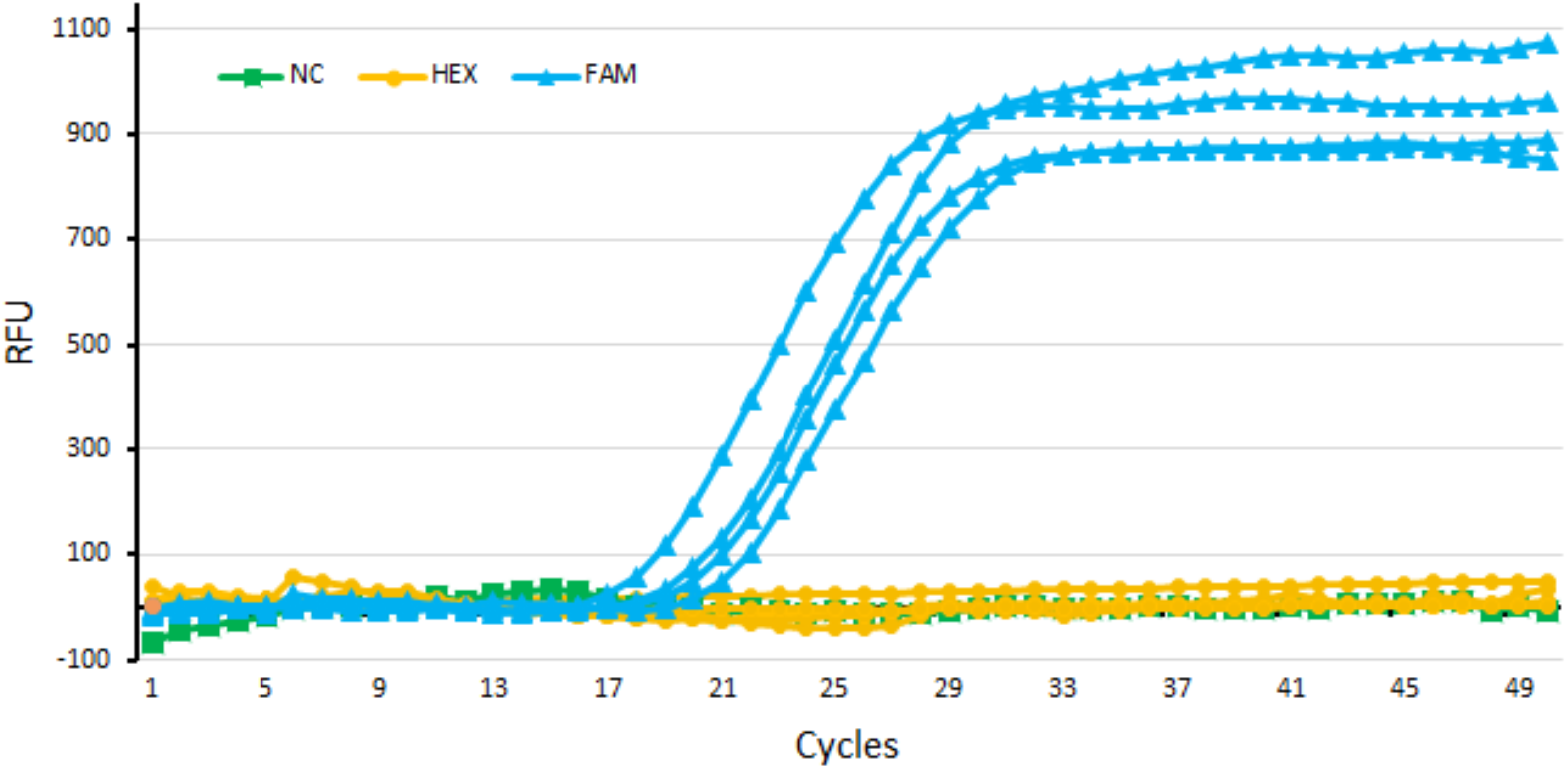
Amplification curves using AddProbe rRT PCR master mix (AddBio) using D614G IN primers and probes including D614 D-FAM and G614 G-HEX probes. Only HEX (blue) showed amplification curves indicating the G614 mutant, but FAM (yellow) did not form amplification curves indicating that wild type D614 is absent. NC (green) is a negative control.

### 3.2. ARMS PCR

The single plex reactions (Panel A) performed for D614 AF and Out R primers showed no PCR products (Figure 4-A-1) that indicates wild type D614 variant is lacking. The second single plex reaction performed using G614 GR and Out F primers, a PCR product of approximately 176 bp indicates G614 mutant (Figure 4-A-2). The third single plex reaction shows PCR products (266 bp) using Out F and Out R primers confirmed that the outer primers worked (Figure 4-A-3). Multiplex ARMS PCR, using all primers in a single tube (Panel B), shows two PCR products with sizes of 176 bp and 266 bp, which indicated G614 mutant and outer PCR products, respectively. The results of ARMS PCR have corresponded with the rRT PCR data.

**Figure 4:**
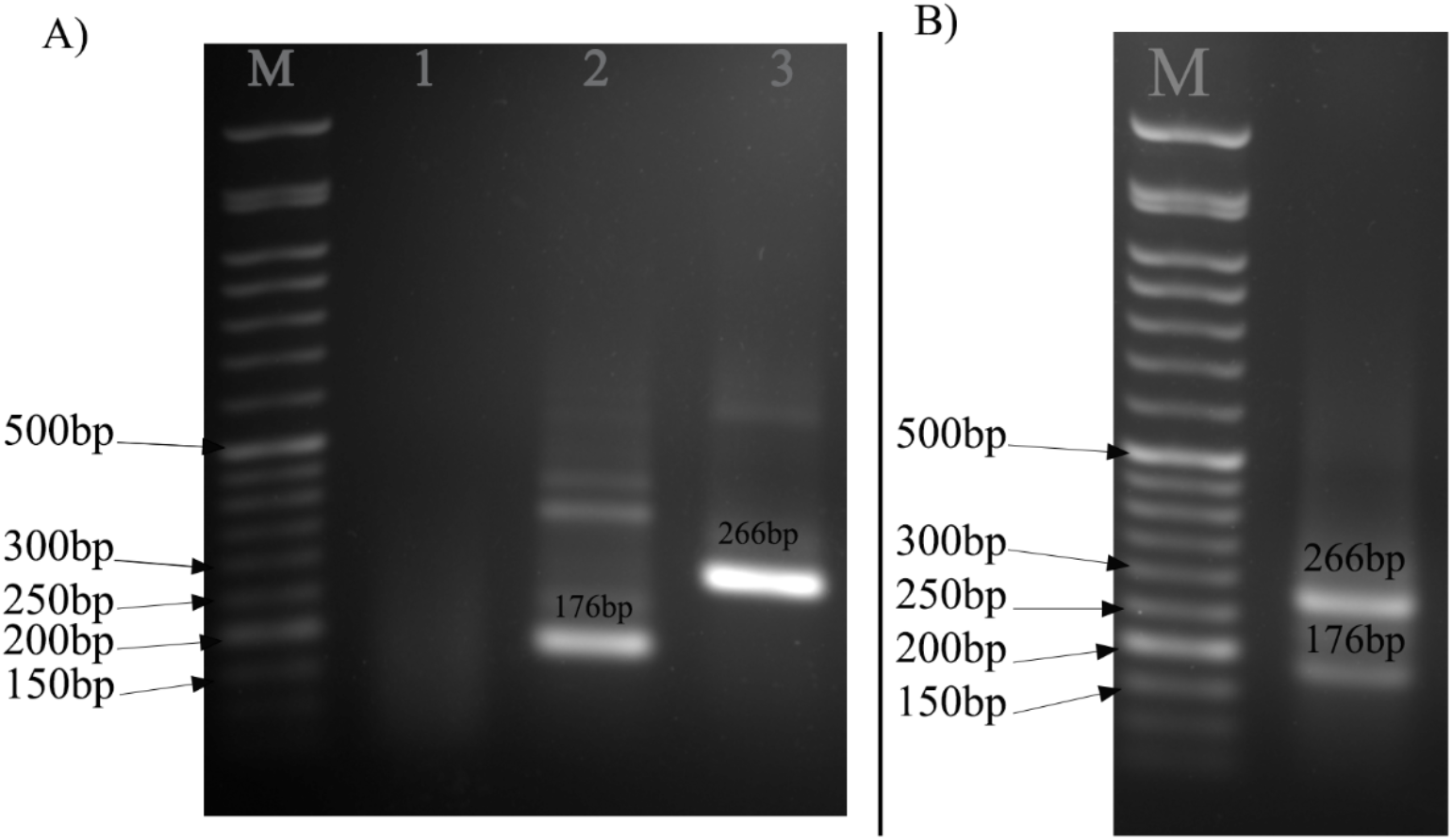
PCR products on 1.5% agarose gel electrophoresis: Addscript RT PCR master mix (AddBio) using ARMS PCR primers. **Panel A:** PCR products using primers separately. Well 1: PCR products using D614 AF and Out R primes. Well 2: PCR products using G614 GR and Out F primers generating 176 bp. Well 3: PCR products using Out F and Out R primers creating 266 bp. **Panel B:** an example of a PCR Product using all primers D614 AF, G614 GR, Out F and Out R in a single tube reaction that created 176 bp (for G614 mutant) and 266 bp (for outer primers).

### 3.2. RFLP

Incubations of PCR products, amplified by D614G Out primers, with BtsCI enzyme, produced no cleavages indicated that only G614 mutants of SARS CoV-2 are prevalent in the region. The enzyme validity was checked by incubations with human cytochrome b PCR products amplified by universal primers as described in section 2.4.3. The results showed that the BtsCI enzyme has cleaved the PCR products of the human cytochrome b (Figure 5).

**Figure 5:**
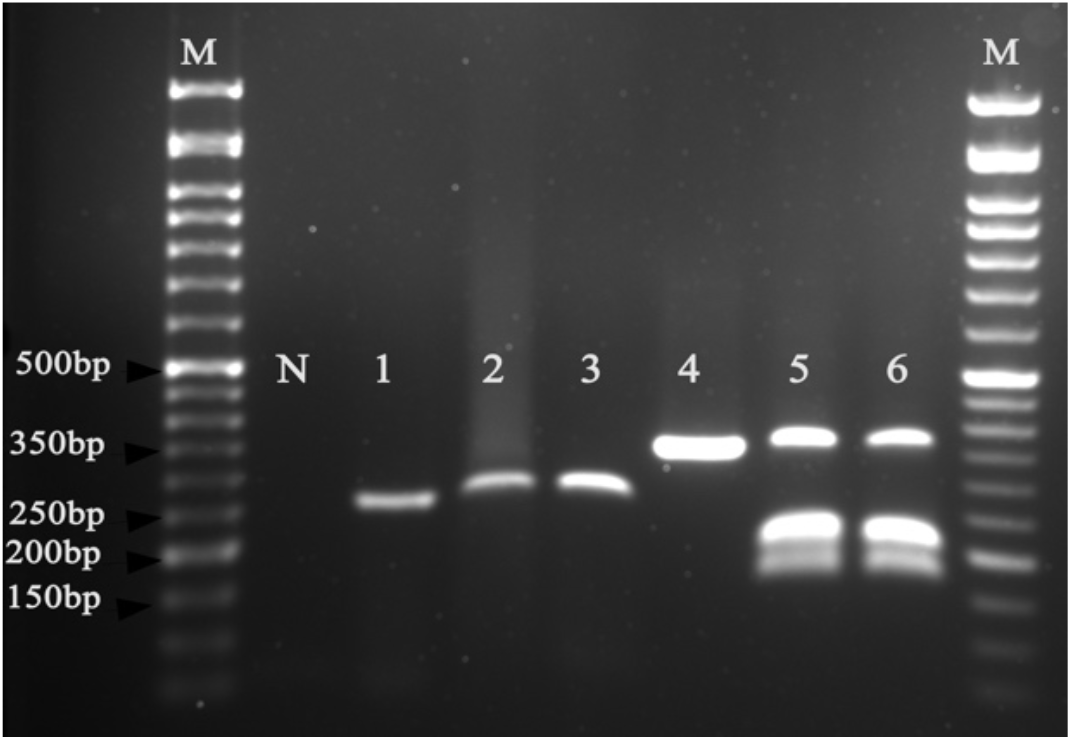
PCR products on 1.5% agarose gel electrophoresis using Addscript RT PCR Master mix (AddBio) and ARMS PCR primers. M= DNA marker 50bp. N= negative control. Lane 1= undigested PCR product amplified by D614G Out primers. Lanes 2 and 3= PCR products amplified by D614G Out primers and incubated with BtsCI enzyme at 50°C. Lane 4= undigested human cytochrome b PCR products amplified by universal primers. Lanes 5 and 6= the human cytochrome b PCR products were cleaved by the BtsCI enzyme.

### 3.3. DNA squences

The DNA sequences results showed only G614 variants as Genbank accession numbers (<u>MW405786</u>, <u>MW405787</u> and <u>MW405788</u>) were released in NCBI online data base.

## 4. Discussions

In the current study, we developed three rapid and inexpensive techniques that can be used to discriminate between D614 and G614 variants. The methods included rRT PCR, ARMS and RFLP. Data obtained by these methods revealed that only G614 mutants are prevalent in the studied region. The results of all of the three methods are corresponding with each other and the DNA sequencing data. We believe our methods are advantageous over SARS CoV-2 whole genome sequences, particularly in developing countries for tracking the D614G mutation and its association with severity, infectivity, and fatality which have been controversial in various studies. It is worth to remember the origin of the virus has not been found. However, WHO has been searching for the viral emergence in Wuhan suggested that the virus may have been arisen from frozen wildlife or SARS CoV-2 related viruses of animals circulated outside China (Wacharapluesadee et al., 2021). The first SARS CoV-2 cases form Wuhan China were considered as an original wild type D614. Thus, future re-emergence of the D614 is not unlikely. Therefore, reasonable diagnostic techniques are required for discovering future D614 outbreaks or tracking previous retro-perspective samples preserved in developing countries. For example in Iraq, a whole genome sequencing has been performed for one sample concluding for discovering G614 mutation that may help understanding the spread of the virus (Al-Rashedi et al., 2021). Therefore, retro-perspective studies for hunting both D614 and G614 are an area of interest in developing countries.

Previous *in vitro* and simulation studies have found that G614 mutation is considered as more infectious variant than the D614 wildtype (Daniloski et al., 2020; Hou et al., 2020; Hu et al., 2020; Korber et al., 2020; Li et al., 2020; Wang et al., 2020). Studies showed that D614G mutation is associated with antigenic epitopes (Gupta et al., 2020; Hernández-Huerta et al., 2020; Islam et al., 2020; Kim et al., 2020; Koyama et al., 2020; Saha et al., 2020; To et al., 2020), antibody neutralizing sensitivity (Goldman et al., 2020;Hu et al., 2020; Klumpp-Thomas et al., 2020; Li et al., 2020; Mansbach et al., 2020), infectivity (Daniloski et al., 2020; van Dorp et al., 2020; Hu et al., 2020; Korber et al., 2020; Li et al., 2020), transmission (van Dorp et al., 2020) and fatality (Hernández-Huerta et al., 2020). Plante *et al* 2020 have suggested that SARS CoV-2 G614 variant has roles in increasing upper respiratory viral loads, transmissions and survival fitness of the virus (Plante et al., 2020). To explore whether this SNV was as a result of either founder effects or viral fitness, caused by random mutation in the viral genome, larger clinical data could be linked with this SNV by comparing SARS CoV-2 disease severity with each D614 or G614 subtypes.

The global vaccines against SARS CoV-2 have been designed using the wildtype D614 virus. One of the most problematic features, which play roles in succeeding vaccines and monoclonal antibodies, is viral SNVs that make the virus resist neutralizing antibodies produced by the vaccines or immune-therapeutics. Therefore, some vaccines or designed antibodies may not be effective against all SARS CoV-2 variants. Thus, identifications of SARS CoV-2 SNVs may help researchers to examine vaccination programs and immunological drugs (Fernández, 2020a, 2020b), before vaccine or drug trial steps.

There have been controversies around the impacts of D614G mutations on vaccine developments. For instance, in a study that used both *in vitro* experiments and structural modelling, G614 has been shown to have no effects on vaccines (McAuley et al., 2020). However, it has been revealed that SARS CoV-2 mutations impact the viral spike antigenic epitopes which are normally recognized and neutralized by antibodies produced by B-lymphocytes (Fernández, 2020b).

A limitation of this study was the absence of the D614 wild type subtype in the area where SARS CoV-2 has been spreading. However, in our study, three assays, as Taqman probes, ARMS, and RFLP all validated each other, in addition to the DNA sequencing data.

Co-segregations of SNVs are also essential for future understanding of the roles of viral pathogenesis and host immune responses against the virus. For instance, identifications of common co-occurred mutations in SARS CoV-2 spike proteins at a particular population would overcome problems of the viral antibody resistance and host cell binding capacity of the virus. By 12 February 2021, the D614G (A23403G) is the most prevalent single nucleotide variations (SNVs) in the world that is about 439,559 sequences (95.5%). Other common mutations in the spike (S) protein, including A222V (C22227T), L81S (C21614T), and S477N (G22992A), are co-occurred with D614G in 100,401 sequences (21.8%), 45,943 sequences (10%) and 23,426 sequences (5.1%), respectively (Covid19 CG data: https://covidcg.org/?tab=group). Moreover, a UK variant, N501Y, has been emerged since October 2020 and currently spreading in the country (Leung et al., 2021) and has become public threat in more than 30 countries (WHO, 2020). Therefore, future work should concentrate on these four mutations to see whether their co-occurrences with D614G are associated with disease severity, infectivity, transmissions, and antibody sensitivity.

Moreover, mutations play roles in zoonotic transmissions from animals to humans and vice versa. A recent study has shown that mammals such as mink harbour SARS CoV-2 mutations, which make the virus resistant to neutralizing antibodies from recovered patients (Oude Munnink et al., 2020). However, minks have naturally infected with both D614 and G614 variants of SARS CoV-2, no evidence shows the impact of G614 mutations in zoonotic outbreaks in this species of mammals. Nonetheless, this suggests that mink and other mammals could have been reservoirs for both D614 and G614 variants that their outbreaks might occur in future. Further studies using large numbers of samples are necessary to discover this effect. Besides, re-infections with SARS CoV-2 are also rare in the world (Goldman et al., 2020; To et al., 2020) and it is also essential to study the roles of mutations including G614 variants in re-infections. Rapid and cost-effective methods are also essential to study the roles of mutations in zoonosis, animals reservoirs and re-infections.

## 5. Conclusions

Up to our best knowledge, this is the first assay developed specifically for exploring SARS CoV-2 D614G mutation using specific primers and probes for detecting D614G mutant by using real-time, conventional PCR and restriction endonuclease. This will help developing countries to conduct further research on this mutation concerning the origin of the virus using previously preserved samples and re-infections occurred between the pandemic waves. Future research should focus on other common co-segregated mutations occurred SARS CoV-2 genome.

## Data Availability

The DNA sequences results showed only G614 variants as Genbank accession numbers (MW405786, MW405787 and MW405788) were released in NCBI online data base.

https://www.ncbi.nlm.nih.gov/nuccore/MW405786

https://www.ncbi.nlm.nih.gov/nuccore/MW405787

https://www.ncbi.nlm.nih.gov/nuccore/MW405788

## Abbreviations

SARS CoV-2: severe acute respiratory syndrome coronavirus 2
D614G: Aspartate 614 mutated to Glycine
ARMS: amplification refractory mutation system RT
RFLP: restriction fragment length polymorphism

## Acknowledgements

We would like to thank Sheikh Hassan Al-Talabani for his great support building our new laboratory, Coronavirus Research and Identification Lab., amid Covid-19 becoming pandemic.

## Authors statements

Conceptualization: SMAA, and SSN; Data curation: SMAA, and SSN; Formal analysis SMAA, and SSN; Investigation: all authors; Methodology: SMAA, and SSN; Resources: all authors; Validation: SMAA, and SSN; Visualization: SMAA, and SSN; Writing - original draft: SMAA, and SSN; Writing - review & editing: all authors.

